# Cannabis Use and Cannabis Use Disorder among U.S. Adults with Psychiatric Disorders: 2001-2002 and 2012-2013

**DOI:** 10.1101/2024.02.29.24303158

**Authors:** Deborah S. Hasin, Zachary L. Mannes, Ofir Livne, David S. Fink, Silvia S. Martins, Malki Stohl, Mark Olfson, Magdalena Cerdá, Katherine M. Keyes, Salomeh Keyhani, Caroline G. Wisell, Julia M. Bujno, Andrew Saxon

**Author notes:** **Corresponding Author:** Deborah Hasin, Ph.D., Columbia University/New York State Psychiatric Institute, 1051 Riverside Drive, Box 123, New York, NY 10032.

## Abstract

**Objective:** Rates of cannabis use disorder (CUD) have increased disproportionately among Veterans Health Administration (VHA) patients with psychiatric disorders, but determining whether such an increase occurred more generally among U.S. adults requires nationally representative data.

**Methods:** Data came from 2001-2002 (n=43,093) and 2012-2013 (n=36,309) national surveys. Outcomes were any past-year non-medical cannabis use, frequent non-medical use (≥3 times weekly), and DSM-IV CUD. Psychiatric disorders included mood, anxiety disorders, antisocial personality disorder, and bipolar I. Logistic regressions were used to generate predicted marginal prevalences of the outcomes for each survey, risk differences calculated, and additive interaction tests determined whether between-survey differences in risk of cannabis outcomes differed between those with and without psychiatric conditions.

**Results:** Cannabis outcome prevalences increased more among those with than without any psychiatric disorder. The difference in prevalence differences included any past-year non-medical cannabis use, 2.45% (95%CI=1.29, 3.62); frequent non-medical cannabis use, 1.58% (95%CI=0.83, 2.33); CUD, 1.40% (95%CI=0.58, 2.21). For each specific disorder, prevalences increased more among those with the disorder, except CUD among those with antisocial personality disorder.

**Conclusions:** In the U.S. general population, rates of cannabis use and CUD increased at a greater rate among adults with psychiatric disorders, similar to findings from VHA patients. These results suggest that although VHA patients are not representative of all U.S. adults, findings from this important patient group can be informative. As U.S. cannabis use continues to expand, greater clinical and policy attention to CUD is needed for adults with psychiatric disorders.

## Introduction

Over the last two decades, US adults’ perception of risk in cannabis use has decreased^1-3^ and cannabis use for medical or recreational purposes has been legalized in most U.S. states^4,5^. Back in 2002, 10.4% of U.S. adults used cannabis and 1.5% had a diagnosis of Cannabis Use Disorder (CUD)^6^. By 2022, rates were sharply higher: 23.0% of U.S. adults had used cannabis in the past year and 6.9% had a past-year CUD diagnosis^7^. Thus, cannabis is one of the most widely-consumed psychoactive substances in the U.S^8^, and CUD is a highly prevalent substance use disorder.

Although some individuals can use cannabis without harm, meta-analysis shows that approximately one-third of those who use cannabis regularly have CUD^9^, and prospective studies show that greater frequency of cannabis use at baseline substantially increases the risk of later CUD^10^. Additional adverse outcomes of cannabis use include psychiatric symptoms and disorders (e.g., anxiety, depression, psychosis)^11-14^. While the direction of the relationship between cannabis use and psychiatric disorders is uncertain, the literature is clear that cannabis use and CUD occur at disproportionately high rates among those with psychiatric disorders.^11,15^ However, little is known about whether the increases over time in the prevalence of cannabis use and CUD are disproportionately greater in individuals with psychiatric disorders than among those without such disorders.

Ideally, determining whether rates of cannabis use and CUD increased disproportionately among those with diagnoses of specific psychiatric disorders would utilize data from nationally representative samples of the general population assessed repeatedly from several years ago to the current time. However, such data do not exist. A study addressing this question used 2005 to 2019 Veterans Health Administration (VHA) national electronic medical record patient data^16^. This study showed that rates of increase in CUD prevalence were substantially greater among patients with psychiatric disorders (e.g., depressive, anxiety, and bipolar disorders) than among patients with no psychiatric disorder.^16^ However, while the VHA is a national healthcare system, VHA patients are more likely to be male, older and socioeconomically disadvantaged than the U.S. adult general population^17^, and thus are not nationally representative. Because of this, findings on CUD prevalence trends in VHA patients may not be generalizable or applicable to others. To address this concern, we identified an additional source of data that was nationally representative, two nationally representative surveys of US adults conducted about 10 years apart. These surveys are the 2001–2002 National Epidemiologic Survey on Alcohol and Related Conditions (NESARC)^18^ and the 2012–2013 National Epidemiologic Survey on Alcohol and Related Conditions–III (NESARC-III)^19^. While these data are older, they enabled us to conduct a study comparing changes in the prevalence of non-medical cannabis use, frequent non-medical cannabis use, and CUD among adults in the US general population between participants with and without psychiatric diagnoses. We examined whether increases over time in non-medical cannabis use, frequent non-medical cannabis use (≥3 times weekly), and DSM-IV CUD were greater in participants with any of a set of common psychiatric disorders than in participants without such disorders, and then examined increases by specific disorders.

## Methods

### Sample and Procedures

NESARC protocols and consent procedures were approved by the institutional review boards at the US Bureau of the Census and the Office of Management and Budget; written consent was provided. The NESARC-III protocols and consent procedures were approved by institutional review boards at the National Institutes of Health and Westat; consent was verbal but recorded electronically, as approved by both institutional review boards. The NESARC^18^ and NESARC-III^19^ used similar multistage probability designs to create samples of adults age ≥18 living in households and group quarters^20^. Each sample was independently ascertained. Both surveys were sponsored by the National Institute on Alcohol Abuse and Alcoholism (NIAAA), who contracted the field work to large survey organizations (NESARC: US Census Bureau; NESARC-III: Westat). The total number of participants analyzed was 79,402 (NESARC n=43,093, NESARC-III n=36,309). Field procedures for both surveys were rigorously overseen by NIAAA staff^21,22^, including sample recruitment, interviewer training (structured home-study, in-class training) and supervision by trained field supervisors, and random respondent callbacks to verify interview data. The methodological similarities of the two surveys have facilitated examination of change over time in numerous important health outcomes^22-25^. The NESARC response rate was 81.0%; the protocol and written consent procedures were approved by US Bureau of the Census and Office of Management and Budget Institutional Review Boards (IRB). The NESARC-III response rate was 60.1%, similar to other contemporaneous nationally representative surveys^26,27^. Sample weights adjusted for selection probabilities and nonresponse. The protocols and consent procedures (verbal, recorded electronically) were approved by IRBs at the National Institutes of Health and Westat.

### Measures

The Alcohol Use Disorder and Associated Disabilities Interview Schedule (AUDADIS), a structured computer-assisted diagnostic interview^11,28-32^, was used to assess non-medical cannabis use, CUD, psychiatric disorders, and sociodemographic characteristics.^20^ Study outcomes consisted of three past-year (past 12 months) cannabis variables: any non-medical use, frequent non-medical use, and DSM-IV cannabis use disorder (CUD). Non-medical use was defined as use without a prescription or other than prescribed, e.g., to get high^21^. Any use was defined as ≥1 time in the past 12 months, and frequent use as ≥3 times weekly. CUD was defined as meeting criteria for DSM-IV cannabis dependence or abuse, as has been reported previously.^22^ These two disorders were combined because their criteria reflect a single condition^33^. Most of the 22 CUD symptom items used in both surveys were identical; examination of the few slight differences showed that they could not account for the large differences in prevalence in the two surveys^21,22^.

The psychiatric disorders analyzed in the present study included depressive disorders (major depressive disorder, dysthymia), anxiety disorders, antisocial personality disorder, and bipolar I disorder. Any anxiety disorder included panic disorder with and without agoraphobia, social phobia, specific phobia, and generalized anxiety disorder. All psychiatric diagnoses excluded substance- and medically-induced disorders. For each individual psychiatric disorder, a binary variable was created to include only those with the disorder and those without *any* of the listed psychiatric disorders. A variable was also created to represent any psychiatric disorder (APD) vs. none.

Multiple versions of the AUDADIS, including those used in the NESARC and NESARC-III, have been subjected to rigorous reliability testing^11,28-32^. Most recently, in a general population sample, test-retest reliability of 12-month cannabis use was substantial (kappa=0.78)^28^, and test-retest reliability of CUD (kappa=0.41) and its dimensional criteria scale (intraclass correlation coefficient=0.70) was fair to substantial in a general population^34^. Validity of CUD was assessed through blind clinician re-appraisal using the semi-structured, clinician-administered Psychiatric Research Interview for Substance and Mental Disorders (PRISM)^35^ in a separate general population sample. AUDADIS/PRISM concordance was moderate for cannabis use disorder (kappa=0.60) and substantial for its dimensional criteria scale (ICC=0.79). Reliability and validity of depressive and anxiety disorder diagnoses was fair to moderate.

Study control covariates included respondent gender (male, female); age (18-29; 30-44; 45-64; ≥65 years); race/ethnicity (Hispanic; Non-Hispanic White and Black; Native American; Asian/Pacific Islander); education (<high school; high school graduate or GED; ≥some college); marital status (married or living together; separated/widowed/divorced; never married); family income ($0-$19,999; $20,000-$34,999; $35,000-$69,999; ≥$70,000).

### Statistical Analysis

NESARC and NESARC-III data sets were concatenated. To evaluate trends between the two surveys, a survey indicator variable was added. We used logistic regressions to model cannabis outcomes as a function of psychiatric disorder, survey, and psychiatric disorder by survey interactions, with control covariates including sociodemographic characteristics (age, race/ethnicity, gender, education, marital status, income) and covariate-by-psychiatric disorder interactions, which allow covariate effects to differ between those with and without psychiatric disorders. For each cannabis outcome, predictions from the logistic models were used to generate predicted marginal prevalences of the cannabis outcomes in each survey (prevalences standardized to the distribution of sociodemographic characteristics of the sample pooled across the two surveys) by psychiatric disorder status (yes/no). Risk differences were calculated for between-survey comparisons (2001-2002 compared with 2012-2013) within each psychiatric disorder group (i.e. those with and without any psychiatric disorder, and then each psychiatric disorder). Interaction on the additive scale tested whether the difference in absolute risk of each cannabis outcome between surveys differed between those with and without each psychiatric disorder (difference in risk differences, or DiD). Interactions on the additive scale were evaluated because they indicate groups with the greatest population-level risk, or changes in risk, and thus can be considered most informative from a public health perspective. To conduct the analyses, SUDAAN v.11.0.4 was used, incorporating survey weights to adjust for the complex sampling design and to yield U.S. adult population-representative estimates. Estimates for risk differences and 95% confidence intervals were obtained using the PRED_EFF option in SUDAAN. Statistical tests were two-tailed, with significance based on p<0.05, as indicated by 95% confidence intervals. For difference effects, a value of 0.0 indicates no difference, so an estimate with a 95% confidence interval that does not include 0.0 is statistically significant at p<0.05.

## Results

Sample characteristics are shown in Table 1. The samples had similar distributions across sociodemographic groups. In 2001-2002, 19.07% (standard error [SE]=0.14) had any psychiatric disorder (APD) as defined for this study, while in 2012-2013, 23.99% (SE=0.38) had APD.

**Table 1.** Characteristics of the two survey samples.

|  | **NESARC**<br>(N=43,093)<br>2001-2002 |  | **NESARC-III**<br>(N=36,309)<br>2012-2013 |  |
| --- | --- | --- | --- | --- |
|  | n | % (SE) | n | % (SE) |
| **Demographics** |  |  |  |  |
| *Sex* |  |  |  |  |
| Men | 18518 | 47.92 (0.15) | 15862 | 48.09 (0.30) |
| Women | 24575 | 52.08 (0.15) | 20447 | 51.91 (0.30) |
| *Age* |  |  |  |  |
| 18-29 | 8666 | 21.80 (0.16) | 8126 | 21.67 (0.36) |
| 30-44 | 13382 | 30.89 (0.16) | 10135 | 25.73 (0.33) |
| 45-64 | 12840 | 31.06 (0.14) | 12242 | 35.03 (0.32) |
| 65+ | 8205 | 16.24 (0.10) | 5806 | 17.57 (0.37) |
| *Race/ethnicity* |  |  |  |  |
| White | 24507 | 70.89 (0.21) | 19194 | 66.19 (0.77) |
| Black | 8245 | 11.07 (0.17) | 7766 | 11.79 (0.66) |
| Native American | 701 | 2.12 (0.09) | 511 | 1.56 (0.12) |
| Asian or Pacific Islander | 1332 | 4.36 (0.05) | 1801 | 5.73 (0.47) |
| Hispanic | 8308 | 11.56 (0.10) | 7037 | 14.74 (0.67) |
| *Annual Family Income* |  |  |  |  |
| \$0-\$19,999 | 12602 | 23.55 (0.15) | 10631 | 22.80 (0.50) |
| \$20,000-\$34,999 | 9401 | 20.19 (0.12) | 7854 | 18.93 (0.35) |
| \$35,000-\$69,999 | 12943 | 32.13 (0.13) | 9562 | 27.21 (0.33) |
| \$70,000+ | 8147 | 24.12 (0.12) | 8262 | 31.06 (0.66) |
| *Education level* |  |  |  |  |
| Less than high school | 7849 | 15.65 (0.13) | 5490 | 13.01 (0.42) |
| High school | 12547 | 29.33 (0.17) | 9799 | 25.78 (0.51) |
| Some college or more | 22697 | 55.02 (0.21) | 21020 | 61.21 (0.76) |
| *Marital status* |  |  |  |  |
| Married/living together | 22081 | 61.62 (0.14) | 16794 | 57.83 (0.51) |
| Separated/widowed/divorced | 11117 | 17.46 (0.11) | 9423 | 19.66 (0.32) |
| Never married | 9895 | 20.92 (0.16) | 10092 | 22.52 (0.44) |
| *Psychiatric disorders* |  |  |  |  |
| Any of the below disorders | 8218 | 19.07 (0.14) | 8870 | 23.99 (0.38) |
| Major Depression | 2422 | 5.47 (0.16) | 3963 | 10.39 (0.25) |
| Any Depressive Disorder (major depression or dysthymia) | 2653 | 5.93 (0.16) | 5357 | 14.05 (0.29) |
| Any Anxiety Disorder | 4886 | 11.37 (0.33) | 4701 | 13.07 (0.24) |
| Antisocial Personality Disorder | 1422 | 3.63 (0.15) | 1600 | 4.33 (0.16) |
| Bipolar 1 Disorder | 1411 | 3.31 (0.13) | 753 | 2.07 (0.10) |

### Increases in cannabis outcomes by Any Psychiatric Disorder (APD; yes vs. no): Table 2

Among those with APD, any non-medical cannabis use increased from 8.03% in 2001-2002 to 15.25% in 2012-2013, an increase of 7.22%. Among those without APD, the increase was from 2.90% in 2001-2002 to 7.67% in 2012-2013, an increase of 4.77%. The difference in prevalence differences (DiD) between the two surveys, 2.45%, indicated a significantly greater increase in any non-medical use among those with APD. Frequent non-medical use among those with APD increased from 2.84% in 2001-2002 to 6.46% in 2012-2013, an increase of 3.62%. Among those without APD, the increase was from 0.68% in 2001-2002 to 2.72% in 2012-2013, an increase of 2.04%. The DiD estimate, 1.58%, indicated a significantly greater increase in frequent non-medical use among those with APD. Past-year CUD increased among those with APD from 3.36% in 2001-2002 to 5.79% in 2012-2013, an increase of 2.43%. Among those without APD, past-year CUD increased from 0.88% in 2001-2002 to 1.91% in 2012-2013, an increase of 1.04%. The DiD estimate, 1.40%, indicated a significantly greater absolute increase in CUD among those with APD.

**Table 2.**
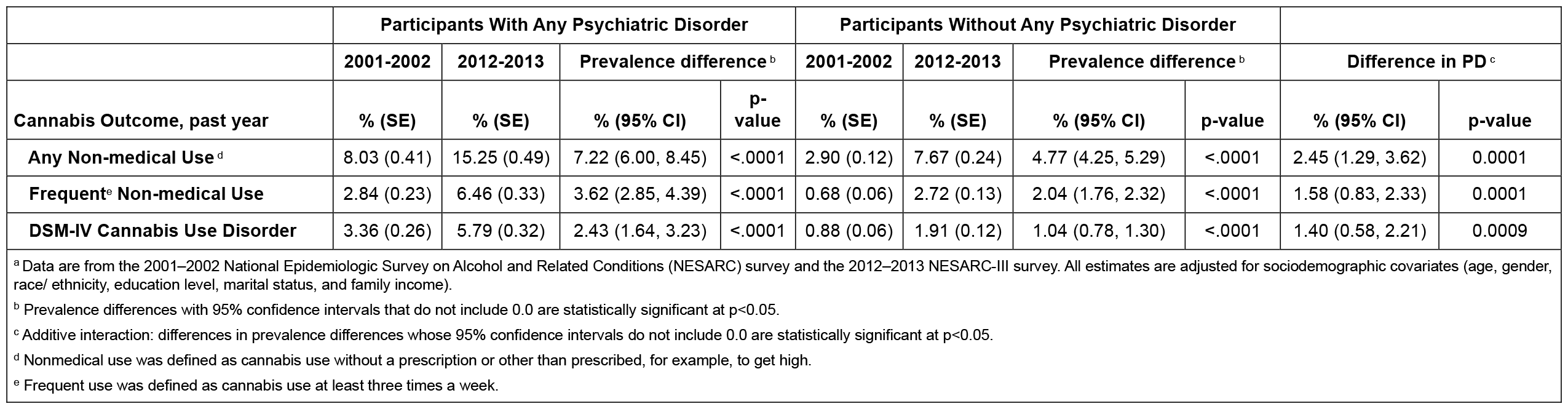
Any non-medical cannabis use, frequent non-medical use, and DSM-IV Cannabis Use Disorder among U.S. adults with and without Any Psychiatric Disorder (APD), 2001–2002 and 2012–2013: Predicted prevalences and between survey and APD group comparisons^a^.

|  | Participants With Any Psychiatric Disorder |  |  |  | Participants Without Any Psychiatric Disorder |  |  |  |  |  |
| --- | --- | --- | --- | --- | --- | --- | --- | --- | --- | --- |
|  | 2001-2002 | 2012-2013 | Prevalence difference <sup>b</sup> |  | 2001-2002 | 2012-2013 | Prevalence difference <sup>b</sup> |  | Difference in PD <sup>c</sup> |  |
| Cannabis Outcome, past year | % (SE) | % (SE) | % (95% CI) | p-value | % (SE) | % (SE) | % (95% CI) | p-value | % (95% CI) | p-value |
| Any Non-medical Use <sup>d</sup> | 8.03 (0.41) | 15.25 (0.49) | 7.22 (6.00, 8.45) | <.0001 | 2.90 (0.12) | 7.67 (0.24) | 4.77 (4.25, 5.29) | <.0001 | 2.45 (1.29, 3.62) | 0.0001 |
| Frequent<sup>e</sup> Non-medical Use | 2.84 (0.23) | 6.46 (0.33) | 3.62 (2.85, 4.39) | <.0001 | 0.68 (0.06) | 2.72 (0.13) | 2.04 (1.76, 2.32) | <.0001 | 1.58 (0.83, 2.33) | 0.0001 |
| DSM-IV Cannabis Use Disorder | 3.36 (0.26) | 5.79 (0.32) | 2.43 (1.64, 3.23) | <.0001 | 0.88 (0.06) | 1.91 (0.12) | 1.04 (0.78, 1.30) | <.0001 | 1.40 (0.58, 2.21) | 0.0009 |

<sup>a</sup> Data are from the 2001–2002 National Epidemiologic Survey on Alcohol and Related Conditions (NESARC) survey and the 2012–2013 NESARC-III survey. All estimates are adjusted for sociodemographic covariates (age, gender, race/ ethnicity, education level, marital status, and family income).
<sup>b</sup> Prevalence differences with 95% confidence intervals that do not include 0.0 are statistically significant at p<0.05.
<sup>c</sup> Additive interaction: differences in prevalence differences whose 95% confidence intervals do not include 0.0 are statistically significant at p<0.05.
<sup>d</sup> Nonmedical use was defined as cannabis use without a prescription or other than prescribed, for example, to get high.
<sup>e</sup> Frequent use was defined as cannabis use at least three times a week.

### Increases in cannabis outcomes by specific psychiatric disorders: Table 3

#### Any non-medical cannabis use

There was a significant increase in the prevalence of any past-year non-medical cannabis use between 2001-2002 and 2012-2013 among those with each psychiatric disorder (from 7.14% to 8.74%) and among those without any psychiatric disorders (from 4.63% to 4.71%). For all diagnoses, the increases were significantly greater among those with each of the psychiatric disorders than among those without the specific disorder (difference in prevalence difference, 2.46% to 4.14%).

**Table 3:** Any non-medical cannabis use, frequent non-medical use, and DSM-IV Cannabis Use Disorder among U.S. adults U.S. adults with and without psychiatric disorders, 2001–2002 and 2012–2013. Predicted prevalences and between survey and psychiatric disorder group comparisons^a^.

|  | With Disorder |  |  |  | Without Disorder |  |  |  |  |  |
| --- | --- | --- | --- | --- | --- | --- | --- | --- | --- | --- |
|  | 2001-2002 | 2012-2013 | Prevalence difference<sup>b</sup> |  | 2001-2002 | 2012-2013 | Prevalence difference<sup>b</sup> |  | Difference in PD<sup>c</sup> |  |
| Psychiatric Disorder | Past Year Any Non-medical Use <sup>d</sup> |  |  |  |  |  |  |  |  |  |
| Major Depression | 6.94 (0.65) | 14.47 (0.61) | 7.52 (5.86, 9.19) | <.0001 | 2.83 (0.11) | 7.49 (0.24) | 4.66 (4.15, 5.17) | <.0001 | 2.86 (1.18, 4.55) | 0.0010 |
| Any Depressive Disorder <sup>f</sup> | 6.98 (0.64) | 15.72 (0.67) | 8.74 (6.99, 10.49) | <.0001 | 2.84 (0.12) | 7.52 (0.24) | 4.68 (4.17, 5.19) | <.0001 | 4.06 (2.33, 5.79) | <.0001 |
| Any Anxiety Disorder | 7.00 (0.50) | 14.35 (0.63) | 7.35 (5.86, 8.83) | <.0001 | 2.80 (0.11) | 7.43 (0.24) | 4.63 (4.12, 5.13) | <.0001 | 2.72 (1.24, 4.20) | 0.0004 |
| Antisocial Personality Disorder | 15.21 (1.15) | 23.41 (1.03) | 8.20 (5.19, 11.21) | <.0001 | 2.87 (0.12) | 7.58 (0.24) | 4.71 (4.20, 5.23) | <.0001 | 3.49 (0.55, 6.43) | 0.0212 |
| Bipolar 1 Disorder | 10.49 (0.86) | 19.27 (1.55) | 8.78 (5.41, 12.15) | <.0001 | 2.82 (0.11) | 7.46 (0.24) | 4.64 (4.14, 5.15) | <.0001 | 4.14 (0.82, 7.46) | 0.0155 |
|  | Past Year Frequent <sup>e</sup> Non-medical Use |  |  |  |  |  |  |  |  |  |
| Major Depression | 1.42 (0.24) | 5.66 (0.44) | 4.24 (3.29, 5.19) | <.0001 | 0.66 (0.05) | 2.64 (0.13) | 1.98 (1.71, 2.26) | <.0001 | 2.26 (1.30, 3.22) | <.0001 |
| Any Depressive Disorder <sup>f</sup> | 1.48 (0.23) | 6.45 (0.45) | 4.96 (3.99, 5.93) | <.0001 | 0.66 (0.05) | 2.65 (0.13) | 1.99 (1.72, 2.27) | <.0001 | 2.97 (2.02, 3.92) | <.0001 |
| Any Anxiety Disorder | 2.36 (0.30) | 6.24 (0.46) | 3.89 (2.87, 4.90) | <.0001 | 0.65 (0.05) | 2.62 (0.13) | 1.97 (1.70, 2.24) | <.0001 | 1.92 (0.92, 2.92) | 0.0002 |
| Antisocial Personality Disorder | 6.29 (0.67) | 11.78 (0.77) | 5.49 (3.53, 7.45) | <.0001 | 0.68 (0.06) | 2.71 (0.13) | 2.03 (1.76, 2.31) | <.0001 | 3.46 (1.51, 5.40) | 0.0006 |
| Bipolar 1 Disorder | 4.25 (0.69) | 9.46 (0.92) | 5.21 (3.02, 7.39) | <.0001 | 0.66 (0.05) | 2.65 (0.13) | 1.99 (1.71, 2.26) | <.0001 | 3.22 (1.04, 5.39) | 0.0042 |
|  | Past Year DSM-IV Cannabis Use Disorder |  |  |  |  |  |  |  |  |  |
| Major Depression | 2.61 (0.37) | 5.60 (0.44) | 2.99 (1.89, 4.10) | <.0001 | 0.85 (0.06) | 1.87 (0.11) | 1.01 (0.76, 1.27) | <.0001 | 1.98 (0.84, 3.12) | 0.0008 |
| Any Depressive Disorder <sup>f</sup> | 2.70 (0.37) | 6.37 (0.48) | 3.67 (2.52, 4.83) | <.0001 | 0.86 (0.06) | 1.87 (0.11) | 1.02 (0.76, 1.27) | <.0001 | 2.66 (1.48, 3.84) | <.0001 |
| Any Anxiety Disorder | 2.95 (0.32) | 5.51 (0.44) | 2.56 (1.53, 3.59) | <.0001 | 0.84 (0.06) | 1.85 (0.11) | 1.00 (0.75, 1.25) | <.0001 | 1.56 (0.53, 2.58) | 0.0034 |
| Antisocial Personality Disorder | 7.15 (0.71) | 9.29 (0.82) | 2.14 (0.05, 4.23) | 0.0459 | 0.88 (0.06) | 1.92 (0.12) | 1.04 (0.78, 1.30) | <.0001 | 1.10 (-0.99, 3.20) | 0.3044 |
| Bipolar 1 Disorder | 5.12 (0.64) | 8.94 (0.99) | 3.82 (1.57, 6.07) | 0.0011 | 0.86 (0.06) | 1.88 (0.12) | 1.02 (0.76, 1.27) | <.0001 | 2.80 (0.52, 5.09) | 0.0173 |

<sup>a</sup> Data are from the 2001–2002 National Epidemiologic Survey on Alcohol and Related Conditions (NESARC) survey and the 2012–2013 NESARC-III survey. All estimates are adjusted for sociodemographic covariates (age, gender, race/ ethnicity, education level, marital status, and family income).
<sup>b</sup> Prevalence differences with 95% confidence intervals that do not include 0.0 are statistically significant at p<0.05.
<sup>c</sup> Additive interaction: differences in prevalence differences whose 95% confidence intervals do not include 0.0 are statistically significant at p<0.05.
<sup>d</sup> Nonmedical use was defined as cannabis use without a prescription or other than prescribed, for example, to get high.
<sup>e</sup> Frequent use was defined as cannabis use at least three times a week.
<sup>f</sup> Includes Major Depressive Disorder or Dysthymia

#### Frequent non-medical cannabis use

There were significant increases in the prevalence of frequent non-medical cannabis use between 2001-2002 and 2012-2013 among both those with each psychiatric disorder (from 3.60 to 5.49%) and those without disorder (from 1.97% to 2.03%). The increase among those with each psychiatric disorder was significantly greater than the increase among those without the disorder (difference in prevalence difference, 1.61% to 3.46%).

#### DSM-IV CUD

The prevalence of CUD increased significantly between 2001-2002 and 2012-2013 among those with each psychiatric disorder (2.14% to 3.82%) and among those without any psychiatric disorder (1.00%-1.04%). With the exception of antisocial personality disorder, the increase among those with each psychiatric disorder was significantly greater than the increase among those without the disorder (difference in prevalence difference, from 1.47% to 2.80%).

## Discussion

This study utilized data from two U.S. nationally representative cross-sectional adult surveys conducted approximately 10 years apart (2001-2002 and 2012-2013) to determine whether increases over time in any past-year non-medical cannabis use, frequent non-medical cannabis use, and cannabis use disorder (CUD) were disproportionately greater among participants with past-year psychiatric disorders versus those without such conditions. The disorders examined were those measured in both surveys, i.e., major depressive disorder and dysthymia, panic disorder with and without agoraphobia, social phobia, specific phobia, generalized anxiety disorder, antisocial personality disorder and bipolar I disorder, as well as a summary variable indicating that one or more of the disorders was present. For any non-medical cannabis use and frequent non-medical cannabis use (≥3 times per week), prevalence increases were greater among those with any psychiatric disorder vs. none, as well as for each of the psychiatric disorders considered separately. For CUD, prevalence increases were greater among those with all psychiatric disorders examined (except antisocial personality disorder), compared to those with none of the psychiatric disorders.

This study was undertaken to explore whether the disproportionate increases in CUD rates among VHA patients with psychiatric disorders, who are more likely to be male, older, White, and have lower income than the U.S. adult general population, would also be found in samples of individuals who were representative of U.S. adult population as a whole. The present results are indeed similar to results found when comparing time trends in the prevalence of CUD between 2005 and 2019 between VHA patients with and without diagnosed psychiatric disorders^16^. In that study, CUD rates also had disproportionately greater increases among patients with psychiatric disorders compared to patients without such disorders. In both studies, the repeated cross-sectional nature of the analysis precluded determining the direction of effect between psychiatric disorders and the cannabis outcomes. However, both sets of results are consistent with an explanation that greater use of cannabis increased the risk for psychiatric symptoms and/or disorders, or that individuals with psychiatric symptoms/disorders increased their use of cannabis to self-medicate the symptoms. To disentangle the direction of effect, fine-grained longitudinal cohort analyses are needed. Such an analysis could potentially be conducted with VHA electronic medical record data. While outside the scope of the present study, this should be done in the future.

Study limitations are noted. NESARC-III are now 12 years old and do not extend into the years of recreational cannabis except for the earliest years in Colorado and Washington. Also, since 2012-2013, the potency of illicitly purchased cannabis has increased, and highly potent legal cannabis products are increasingly available. If another NESARC survey had been conducted around 2020-2022, a similar analysis would reflect more recent changes, but no NESARC surveys were conducted after 2012-2013, precluding such work. Post-traumatic stress disorder (PTSD) was not included because it was not assessed in the 2001-2002 NESARC. Psychotic-spectrum disorders were not included because (a) they were not assessed in detail in the NESARC surveys and (b) an analysis of the relationship of CUD to a self-reported psychosis variable in NESARC surveys has already been reported^36^. DSM-5 CUD diagnoses were not analyzed because not all DSM-5 CUD criteria were not assessed in the 2001-2002 NESARC Depressive and anxiety disorders were diagnosed using DSM-IV criteria in NESARC and DSM-5 criteria in NESARC-III. While rates of major depressive disorder were higher in NESARC-III than in NESARC (Table 1), the minimal changes in the diagnostic criteria did not appear to account for such increases^37^. We note that changing diagnostic criteria are also issues in long-term trend studies using VHA data, which transitioned from ICD-9-CM to ICD-10-CM in 2015. However, despite the changes in nomenclatures across both studies, their findings on disproportionate increases in cannabis outcomes among individuals with psychiatric disorders were consistent. Additionally, all NESARC and NESARC-III data were based on self-report and biological measures were not used. Also, similar to other national surveys, some population segments were not covered in either survey (e.g., homeless and incarcerated individuals), potentially leading to underestimation. Finally, cannabis use variables could have appeared to increase due to reductions in stigma leading to greater reporting over time. However, our findings that urine toxicology tests positive for cannabis in VHA patients also increased over time^38^ suggests that the increases in prevalence of the self-reported cannabis measures in both datasets were not entirely due to reduced stigma of cannabis use.^38^

In conclusion, this study adds to evidence that the adult prevalences of any past-year non-medical use, frequent non-medical use and CUD are increasing faster among individuals with psychiatric disorders than among individuals without psychiatric disorders. The consistency of findings between the older data from representative samples of adults in the general population with the more recent findings on data across among veteran patients treated at the VHA supports the validity of each, and suggests that information gleaned from VHA data may have applicability and utility to patient groups beyond those treated by the VHA. At this stage, factors contributing to the disproportionate increases among those with psychiatric disorders should be sought. Possibilities include changing attitudes towards the efficacy of cannabis to treat psychiatric symptoms such as insomnia, anxiety or chronic pain (which is highly correlated with psychiatric conditions). Another potential contributor consists of the declines in availability of prescribed medications formerly used to treat such symptoms, e.g., opioids, sedatives/hypnotics and anxiolytics, causing a shift to cannabis use and among a vulnerable subset, development of CUD. The findings also provide additional evidence that clinician and policymaker attention to the increasing risks of non-medical cannabis use and CUD among those with psychiatric problems is needed.

## Data Availability

The data is not publicly available.

## Notes

### Competing Interest Statement

Dr. Hasin receives support from Syneos Health for an unrelated project. Dr. Saxon has received consulting fees from Indivior, travel support from Alkermes, research support from MedicaSafe, and royalties from UpTo-Date. Dr. Keyes has served as an expert witness in litigation. The other authors report no financial relationships with commercial interests.

### Funding Statement

This study was supported by NIDA grant R01DA048860, the New York State Psychiatric Institute, and the VA Centers of Excellence in Substance Addiction Treatment and Education.

### Author Declarations

IRB of United States Bureau of the Census gave ethical approval for this work. IRB of United States Office of Management and Budget gave ethical approval for this work. IRB of National Institute of Health gave ethical approval for this work. IRB of Westat gave ethical approval for this work.

